# The impact of SGLT-2 Inhibitors on the Risk of Atrial Fibrillation in Heart Failure

**DOI:** 10.1101/2025.01.21.25320931

**Authors:** Thomas Fretz, Onyedika Ilonze, Tanyanan Tanawuttiwat, Mithilesh K. Das

## Abstract

**Background:** Heart failure (HF) and atrial fibrillation (AF) are closely linked, each exacerbating the other. While sodium-glucose cotransporter-2 inhibitors (SGLT2i) are known to provide substantial benefits in HF management, their effect on AF incidence in this population is not well-defined.

**Objective:** This study aims to assess the effect of SGLT2i therapy on the development of new-onset AF in patients with HF.

**Methods:** This retrospective analysis included all patients hospitalized with a primary diagnosis of HF and no prior diagnosis of AF over a 3-year period. The primary outcome was the occurrence of new-onset AF within 12 months following HF hospitalization.

**Results:** Of 3,953 patients 720 (18.2%) developed AF within one year. SGLT2i use was associated with significantly lower risk of AF (*HR 0.69, 95% CI:0.55-0.87; p=0.002*) across all HF types-reduced, mid-range, and preserved ejection fraction. Kaplan-Meier survival analysis revealed significantly reduced AF-free survival in patients not on SGLT2i therapy, compared to patients on SGLT2i therapy, across subgroups categorized by diabetes, hypertension, coronary artery disease, age ≥65 years, and BMI ≥30Kg/m^2^ (p <0.05 for all). Among those who developed AF, SGLT2i use had significantly lower incidence of AF during follow-up as compared to when not used (12.1% vs. 19.5%, p<0.001). SGLT2 use also delayed onset of AF compared to those not treated with an SGLT2i (339 ± 80 days vs. 317 ± 106 days; *p<0.001*).

**Conclusions:** The use of SGLT2i therapy is associated with a significantly lower risk of the developing AF following hospitalization for HF.

## Introduction

Atrial fibrillation (AF) is the most common arrhythmia in adults and is associated with increased risk of stroke, heart failure (HF), and reduced quality of life. Conversely, HF also increases the risk of AF.^1,2^ Although a causal relationship has not been definitively established, AF and HF frequently co-occur, with AF prevalence estimated at around 25% in all HF patients and up to 50% in those with severe disease^3^ Stroke is a well-known complication of AF and associated with significant morbidity and mortality, but HF is actually the most common complication of AF, with a 4-fold increase in mortality risk in AF patients who develop HF compared to those who experience stroke.^4^ Thus, the development of AF in patients with HF represents a major source of morbidity and mortality.^5–7^

Sodium-glucose cotransporter-2 inhibitors (SGLT2i) are recommended pharmacotherapy of HF, across all ejection fractions (EFs).^8^ SGLT2i reduce HF-related hospitalizations and demonstrate a cardiovascular mortality benefit in patients with HF, cardiovascular disease, and diabetes mellitus (DM).^9^ Meta-analysis and retrospective studies suggest SGLT2i may reduce the incidence or recurrence of AF, and this effect appears consistent regardless of DM or HF status in subgroup analysis.^10–13^ Additionally, recent evidence indicates that SGLT2i use in patients with DM is associated with a lower risk of arrhythmia recurrence after AF ablation.^14,15^ However, to the best of our knowledge, no study has specifically assessed the efficacy of SGLT2i in preventing new-onset AF in patients hospitalized with HF with and without DM. This study aims to characterize the effect of SGLT2i therapy on the incidence of new-onset AF in patients following hospitalization for HF and to evaluate whether SGLT2i therapy delays the onsets of AF in this population.

## Methods

The data analyzed in this study was obtained from the IU Health electronic medical record, encompassing outpatient and specialty-care clinics. Patients aged ≥18 years who were admitted to an IU Health hospitals with a primary diagnosis of HF and without a prior diagnosis of AF between March 2021, and April 2023 were identified using International Classification of Diseases-10th Revision (ICD-10) codes. Patients were included in the study if they either developed AF at a later encounter or had an outpatient visit with a primary care provider or cardiologist at least one year after hospitalization, and if they had an echocardiogram within one year prior to admission. In patients with multiple echocardiograms, the one closest in time to the date of admission was use. Demographic, comorbidity, and medication information were collected from the medical record at the time of discharge, including prescriptions of SGLT2I-i, beta-blockers, mineralocorticoid receptor antagonists, renin-angiotensin system (RAS) inhibitors (angiotensin-converting enzyme [ACE] inhibitors, angiotensin receptor blockers [ARBs], angiotensin receptor-neprilysin inhibitors [ARNIs]). HF classification, comorbidities, and the subsequent development of AF were assessed, with HF classified according to the American College of Cardiology’s criteria based on left ventricular EF. The patient population was divided into two groups based on SGLT2i use. The primary outcome of new diagnosis of AF was analyzed over a one-year period.

Data were analyzed and interpreted by the authors, who affirmed the accuracy and completeness of the data. Exemption from Institutional Review Board (IRB) approval was obtained from the Indiana University given the deidentified nature of the data. These study findings are reported in accordance with the Strengthening the Reporting of Observational Studies in Epidemiology (STROBE) guideline for cohort studies.

### Statistical analysis

The patient population was divided into 2 groups according to their use of SGLT2I inhibitors. Continuous variables are presented as mean ± SD and analyzed using Student’s *t*-tests for normally distributed data and Mann-Whitney *U* tests for non-normally distributed data. Comparisons among more than 2 groups were performed with one-way analysis of variance. Categorical variables were compared by using the chi-square test or Fisher’s exact test. Outcomes were reported as 1-year Kaplan-Meier event rates for AF recurrence and analyzed with Cox proportional hazards regression models, stratified by SGLT2i therapy. A *p-* value <0.05 was considered statistically significant. All analyses were performed by using SPSS statistical software (IBM) version 29.0.

## Results

A total of 3,953 patients were identified and included in our study. Of these, 720 (18.2%) patients developed AF during 1-year follow up. The study population included 880 patients (22.3%) with heart failure with reduced EF (HFrEF), 533 (13.5%) with heart failure with mid-range EF (HFmrEF) and 2,540 (64.3%) with heart failure with preserved EF (HFpEF). New-onset AF was observed in 207 (23.5%) in HFrEF, 107 (20.1%) in HFmEF and 406 (16.1%) in HFpEF patients. All patients received guideline-directed medical therapy (GDMT) as tolerated. **Table 1** lists patient baseline characteristics, subtype of HF, and medication therapy.

**Table 1.** Comparison of patient demographics with and without progression of atrial fibrillation. AF = atrial fibrillation; BMI = body-mass index; LVEF = left ventricular ejection fraction; HFrEF = HF with reduced ejection fraction; HFmrEF = HF with mildly reduced ejection fraction; HFpEF = HF with preserved ejection fraction; COPD = chronic obstructive pulmonary disease; TIA = transient ischemic attack; MRA = mineralocorticoid receptor antagonist; RAS = renin-angiotensin system; SGLT2i = sodium-glucose cotransporter-2 inhibitor; GLP-1 = glucagon-like peptide-1

|  | **No progression to AF**<br>**(n = 3,233)** | **Progression to AF**<br>**(n = 720)** | **P value** |
| --- | --- | --- | --- |
| **Patient characteristics** |  |  |  |
| Age | 65.1 ± 13.2 | 70.9 ± 13.2 | 0.03 |
| Male | 1433 (44.3%) | 364 (50.6%) | <0.001 |
| BMI | 33.7 ± 15.7 | 32.1 ± 13.2 | <0.001 |
| LVEF | 52.5 ± 15.2 | 49.2 ± 13.2 | <0.001 |
| HFrEF | 673 (76.5%) | 207 (23.5%) | <0.001 |
| HFmrEF | 426 (80.0%) | 107 (20.1%) | <0.001 |
| HFpEF | 2134 (84.0%) | 406 (16.0%) | <0.001 |
| Hypertension | 2887 (89.3%) | 503 (69.9%) | <0.001 |
| Dyslipidemia | 2593 (80.2%) | 624 (86.7%) | <0.001 |
| Coronary Artery Disease | 1927 (59.6%) | 487 (67.7%) | <0.001 |
| Diabetes Mellitus | 1875 (58.0%) | 425 (59.0%) | 0.321 |
| Obstructive sleep apnea | 1235 (38.2%) | 290 (40.3%) | 0.160 |
| Chronic kidney disease | 1614 (49.9%) | 473 (65.7%) | <0.001 |
| COPD | 1317 (40.7%) | 354 (49.2%) | <0.001 |
| Prior stroke/TIA | 521 (16.1%) | 106 (14.7%) | 0.193 |
| **Medication therapy** |  |  |  |
| Beta-blocker | 1594 (49.3%) | 335 (46.5%) | 0.096 |
| MRA | 984 (30.4%) | 182 (25.3%) | 0.003 |
| RAS inhibitor | 2315 (71.6%) | 467 (64.9%) | <0.001 |
| SGLT2i | 633 (19.5%) | 87 (12.1) | <0.001 |
| GLP-1 agonist | 445 (13.8%) | 75 (10.4%) | 0.008 |
| Values are n (%) or mean ± SD unless otherwise indicated. |  |  |  |

AF was detected in 87 of 720 (12.1%) patients prescribed SGLT2i therapy compared to 633 of 3249 (19.5%) patients not receiving SGLT2i therapy (P<0.001). This reduction in AF incidence was statistically significant across all HF subtypes and in patients with or without diabetes mellitus (DM). Among patients who developed AF, the onset was delayed in those treated with SGLT2i therapy (338.9 ± 79.7 days vs. 317.5 ± 106 days, p < 0.001) **(Table 2)**. Univariable predictors of AF were male sex, absence of hypertension, hyperlipidemia, chronic kidney disease, chronic obstructive pulmonary disease (COPD), lower EF and SGLT2i use **(Table 3)**. Multivariable predictors of AF were age, ≥65 years, male sex, absence of hypertension, hyperlipidemia, chronic kidney disease, COPD and SGLT2i use **(Table 4)**.

**Table 2.** Patient characteristics who developed atrial fibrillation during follow-up by baseline SGLT2i status. AF = atrial fibrillation; BMI = body-mass index; LVEF = left ventricular ejection fraction; HFrEF = HF with reduced ejection fraction; HFmrEF = HF with mildly reduced ejection fraction; HFpEF = HF with preserved ejection fraction; COPD = chronic obstructive pulmonary disease; TIA = transient ischemic attack; MRA = mineralocorticoid receptor antagonist; RAS = renin-angiotensin system; SGLT2i = sodium-glucose cotransporter-2 inhibitor; GLP-1 = glucagon-like peptide-1

|  | **No SGLT2i**<br>**(n = 633 of 3249, 19.5%)** | **SGLT2i use**<br>**(n = 87 of 720, 12.1%)** | **P value** |
| --- | --- | --- | --- |
| **Age** | 67.1 ± 14.9 | 61.6 ± 13.2 | <0.001 |
| **BMI** | 34.4 ± 9.4 | 33.2 ± 16.3 | 0.96 |
| **AF occurrence from discharge in days** | 317.5 ± 106 | 338.9 ± 79.7 | <0.001 |
| **Male** | 1397(43%) | 400(10.1%) | <0.001 |
| **Female** | 1800 (83.5%) | 356 (16.5%) | 0.001 |
| **Coronary artery disease** | 1927(88.7%) | 487 (12.3%) | <0.001 |
| **Hypertension** | 2780 (82%) | 610 (18.0%) | <0.001 |
| **Diabetes** | 1741 (75.7%) | 559 (24.3%) | <0.001 |
| **Hyperlipidemia** | 2620 (81.4%) | 597 (18.6%) | 0.005 |
| **Obstructive sleep apnea** | 1219 (79.9%) | 306 (20.1%) | 0.002 |
| **COPD** | 1420 (85%) | 251 (15.0%) | <0.001 |
| **Prior stroke/TIA** | 511 (81.5%) | 116 (18.5%) | 0.329 |
| **Chronic kidney disease** | 1728 (82.8%) | 359 (17.2%) | 0.155 |
| Values are n (%) or mean ± SD unless otherwise indicated. |  |  |  |

**Table 3.** Univariable predictors of atrial fibrillation.

|  | RR | 95.0% CI |  | P value |
| --- | --- | --- | --- | --- |
| **Male sex** | **1.26** | **1.058** | **1.502** | **0.010** |
| AGE ≥ 65 | 1.088 | 0.81 | 1.463 | 0.575 |
| **Ejection Fraction** | **0.202** | **0.054** | **0.753** | **0.017** |
| Coronary artery disease | 1.012 | 0.831 | 1.231 | 0.908 |
| **Hypertension** | **0.275** | **0.226** | **0.336** | **<.001** |
| **Dyslipidemia** | **1.694** | **1.276** | **2.249** | **<.001** |
| **Chronic kidney disease** | **1.7** | **1.405** | **2.057** | **<.001** |
| **COPD** | **1.396** | **1.175** | **1.66** | **<.001** |
| BMI | 1 | 0.992 | 1.008 | 0.974 |
| Beta blocker | 0.901 | 0.758 | 1.07 | 0.235 |
| MRA | 0.843 | 0.687 | 1.034 | 0.100 |
| **SGLT2i** | **0.612** | **0.463** | **0.809** | **<.001** |
| GLP-1 agonist | 0.813 | 0.597 | 1.107 | 0.188 |
| **RAS inhibitor** | **0.804** | **0.664** | **0.974** | **0.026** |
| Obstructive sleep apnea | 1.157 | 0.953 | 1.404 | 0.141 |
| Diabetes mellitus | 1.141 | 0.944 | 1.38 | 0.172 |
| Prior stroke/TIA | 0.877 | 0.69 | 1.116 | 0.287 |
| RR = relative risk; CI = confidence interval; other abbreviations as given in Table 1. |  |  |  |  |

**Table 4.** Multivariable predictors of atrial fibrillation.

|  | RR | 95.0% CI |  | p value |
| --- | --- | --- | --- | --- |
| Male sex | 1.352 | 1.166 | 1.567 | <.001 |
| Age ≥ 65 | 1.722 | 1.46 | 2.03 | <.001 |
| Hypertension | 0.245 | 0.207 | 0.291 | <.001 |
| Hyperlipidemia | 1.767 | 1.407 | 2.22 | <.001 |
| Chronic kidney disease | 1.761 | 1.501 | 2.066 | <.001 |
| COPD | 1.312 | 1.131 | 1.523 | <.001 |
| GLP-1 agonist | 0.908 | 0.707 | 1.167 | 0.453 |
| RAS inhibitor | 0.882 | 0.752 | 1.034 | 0.121 |
| Obstructive sleep apnea | 1.091 | 0.935 | 1.273 | 0.267 |
| SGLT2i | 0.667 | 0.527 | 0.845 | <.001 |

Abbreviations as given in Table 1.

Kaplan Meier (KM) survival analysis showed significantly lower AF-free survival in patients with HFrEF compared to HFpEF (*p< 0.001*) and HFmrEF compared to HFpEF (*p=0.019*) but not between HFpEF and HFmrEF (*p=0.14*) **(Figure 1)**. Patients without SGLT2i therapy had significantly lower AF-free survival compared to patients on SGLT2i therapy. **(Figure 2)**. Similar response was consistent across all HF groups (HFrEF, HFmEF and HFpEF, p <0.001 for each) **(Figures 3A-C)**. In diabetic patients, as well as those with hypertension and coronary artery disease, AF-free survival was significantly lower without SGLT2i therapy (**Figure 4-6)**. Kaplan-Meier survival analysis revealed significantly reduced AF-free survival in patients not on SGLT2i therapy compared to those who were on SGLT2i therapy, irrespective of whether patients were ≥65 years old or <65 years old, and regardless of whether their BMI was ≥30 Kg/m2 or <30 Kg/m2. **(Figure 7-8)**. Additionally, patients with chronic kidney disease had significantly lower AF-free survival in patients without SGLT2i therapy as compared to those on SGLT2i therapy.

**Figure 1.**
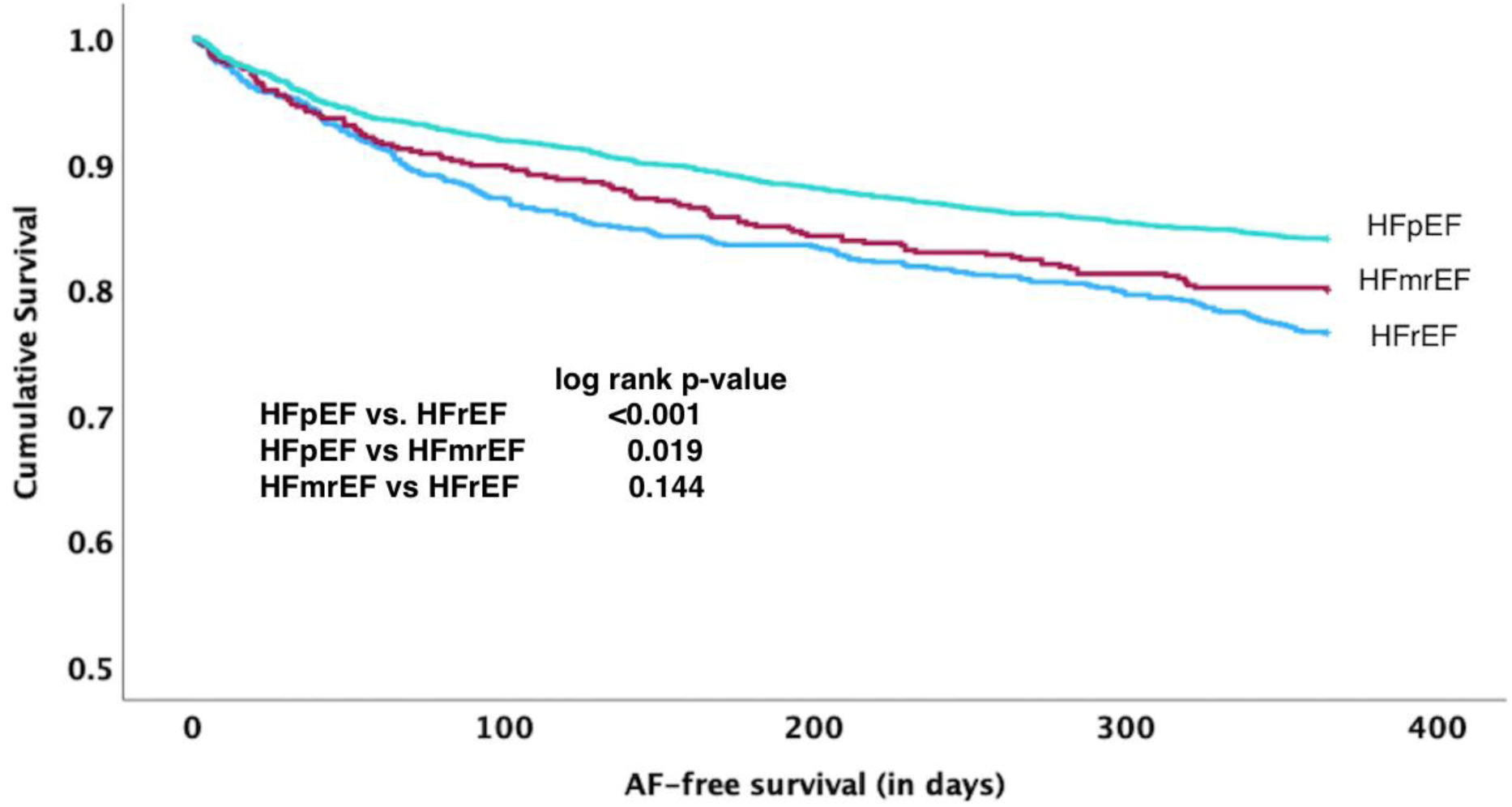
Kaplan-Meier analysis of the freedom from AF recurrence after the HF hospitalization based on HF subtype. During the 12-month follow-up period, AF recurrence was detected in 207 of 880 (23.5%) patients with HFrEF, 107 of 533 (20.1%) of patients with HFmrEF, and 406 of 2540 (16.0%) of patients with HFpEF. The incidence of AF was statistically significant between HFpEF and HFrEF (log-rank test, *P* < 0.001) and between HFpEF and HFmrEF (log-rank test, P = 0.019), but not between HFrEF and HFmrEF (log-rank test, P = 0.144). AF = atrial fibrillation; HFpEF = HF with preserved ejection fraction; HFmrEF = HF with mildly reduced ejection fraction; HFrEF = HF with reduced ejection fraction.

**Figure 2.**
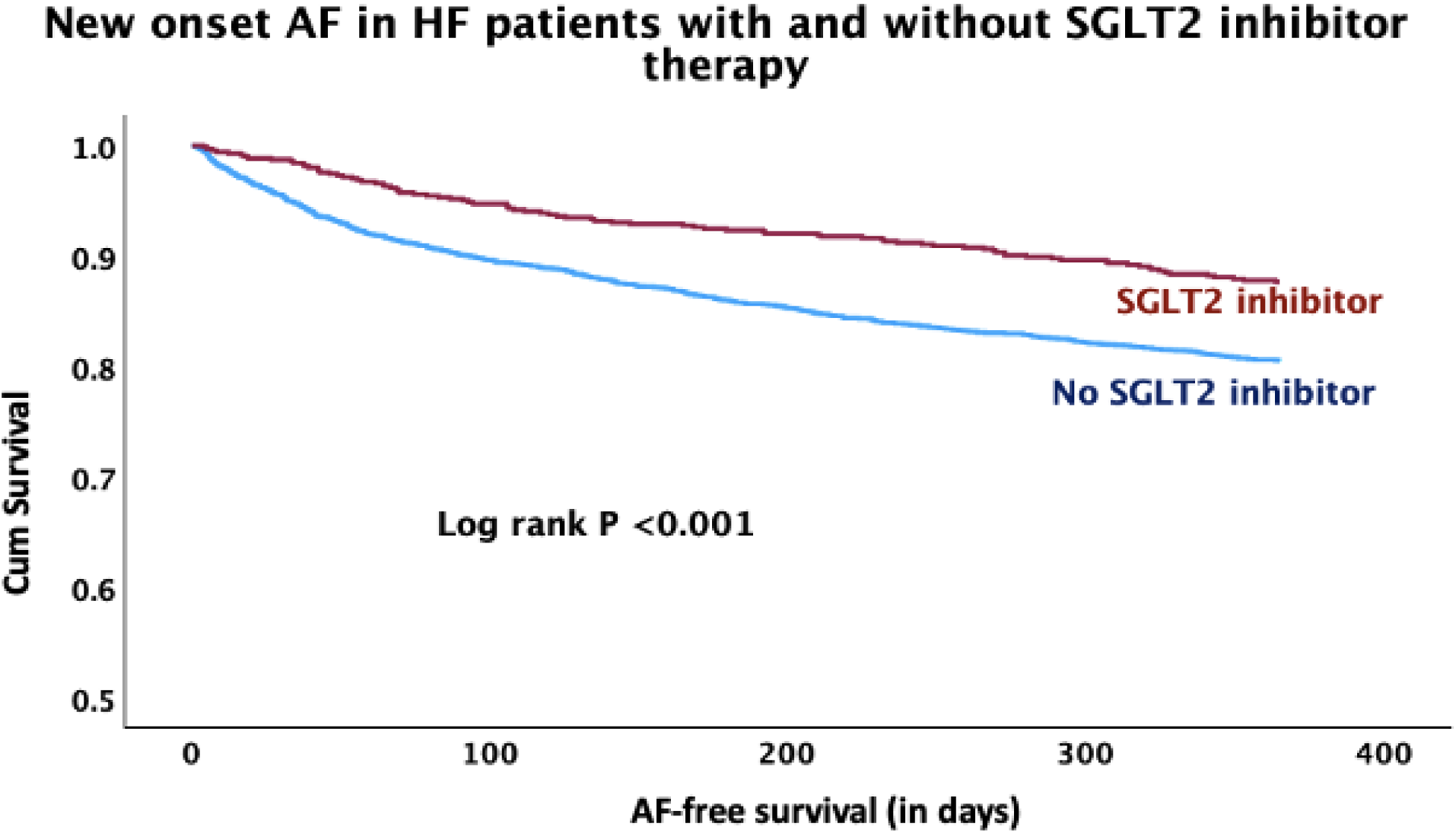
Kaplan-Meier Analysis of the Freedom from AF Following HF Hospitalization. Kaplan-Meier analysis of the freedom from AF recurrence after the HF hospitalization based on SGLT2i therapy. During the 12-month follow-up period, AF recurrence was detected in 87 of 720 (12.1%) patients prescribed SGLT2is compared to 633 of 3250 (19.5%) patients who were not prescribed SGLT2is (log-rank test, *P* < 0.001). AF = atrial fibrillation; SGLT2i = sodium-glucose cotransporter-2 inhibitor.

**Figure 3A.**
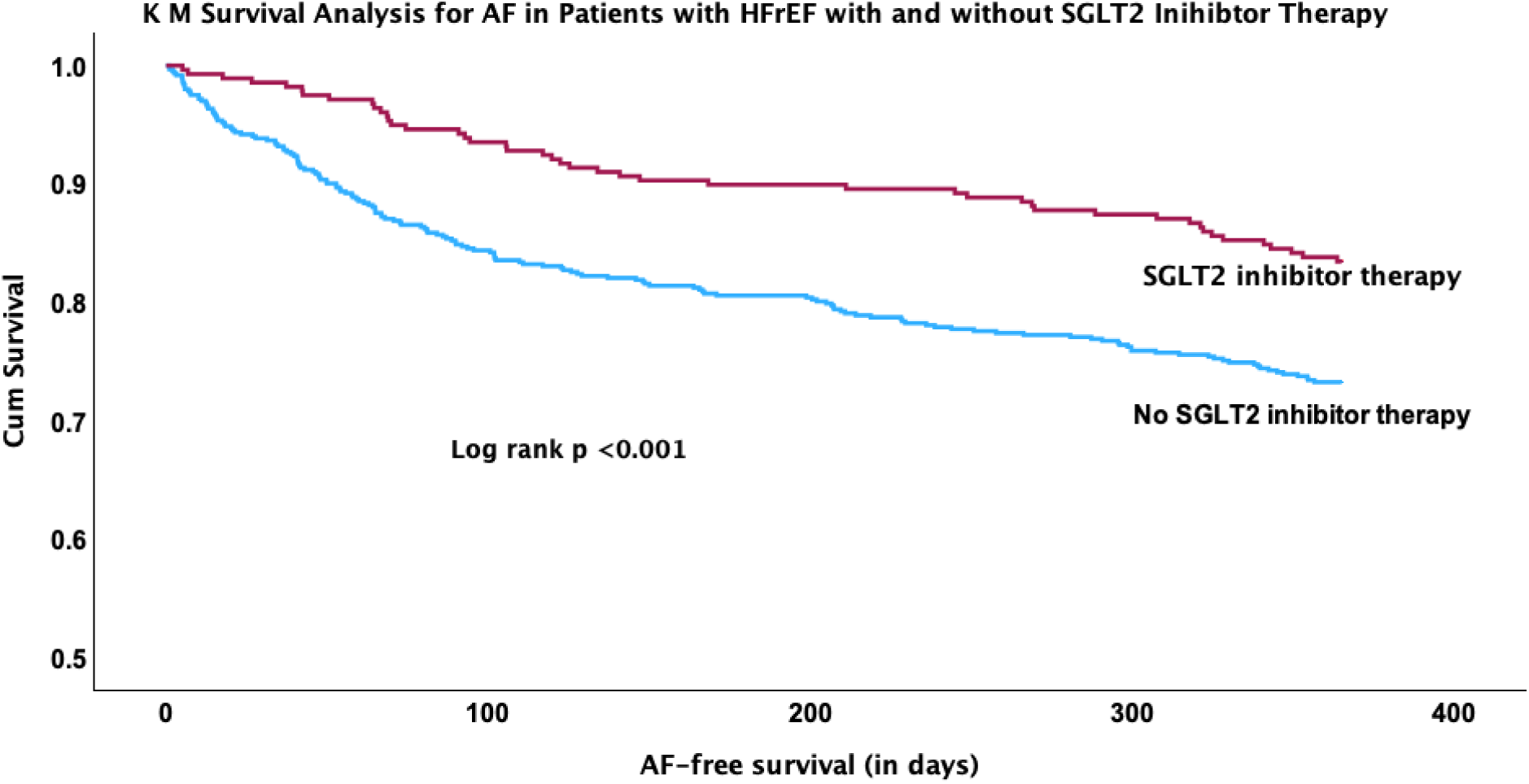
Kaplan-Meier analysis of the freedom from AF following HF hospitalization for HF with reduced Ejection Fraction (HFrEF) in patients with and without SGLT2i therapy. Kaplan-Meier analysis of the freedom from AF recurrence after the hospitalization for HFrEF based on SGLT2i therapy. AF = atrial fibrillation; SGLT2i = sodium-glucose cotransporter-2 inhibitor.

**Figure 3B.**
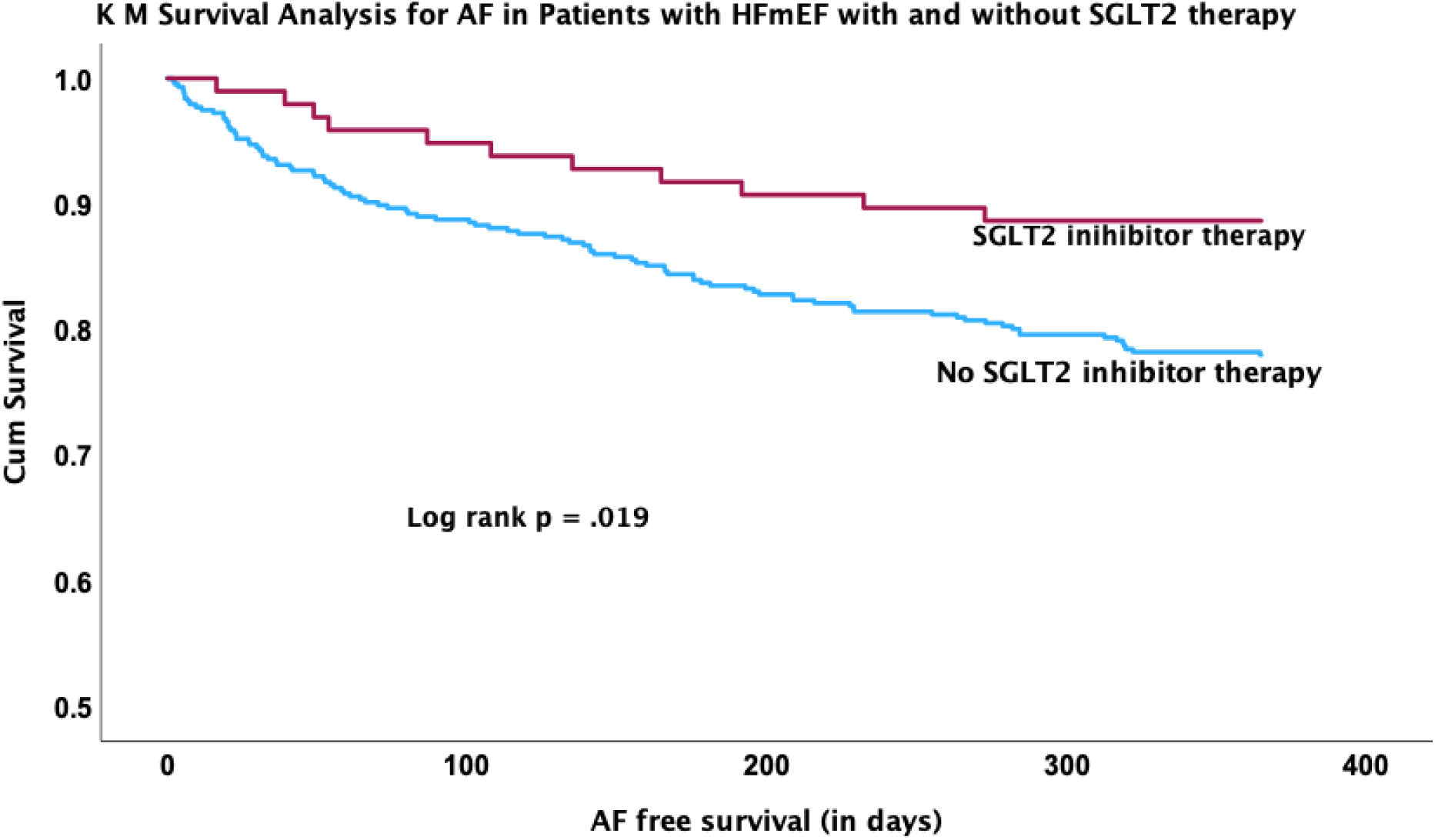
Kaplan-Meier analysis of the freedom from AF following HF hospitalization for HF with medium-range Ejection Fraction (HFmEF) in patients with and without SGLT2i therapy. Kaplan-Meier analysis of the freedom from AF recurrence after the hospitalization for HFrEF based on SGLT2i therapy. AF = atrial fibrillation; SGLT2i = sodium-glucose cotransporter-2 inhibitor.

**Figure 3C.**
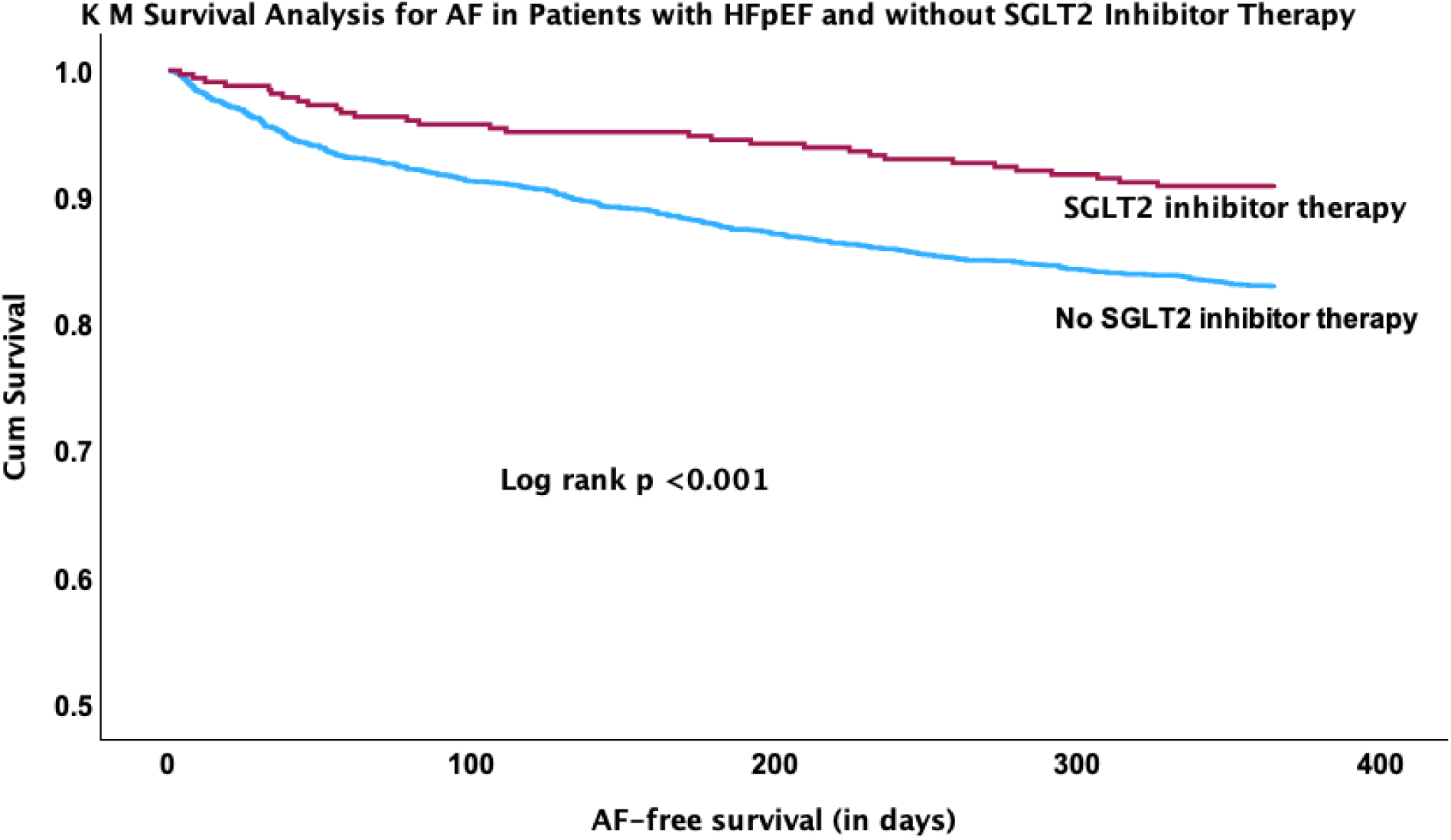
Kaplan-Meier analysis of the freedom from AF following HF hospitalization for HF with preserved Ejection Fraction (HFpEF) in patients with and without SGLT2i therapy. Kaplan-Meier analysis of the freedom from AF recurrence after the hospitalization for HFrEF based on SGLT2i therapy. AF = atrial fibrillation; SGLT2i = sodium-glucose cotransporter-2 inhibitor

**Figure 4.**
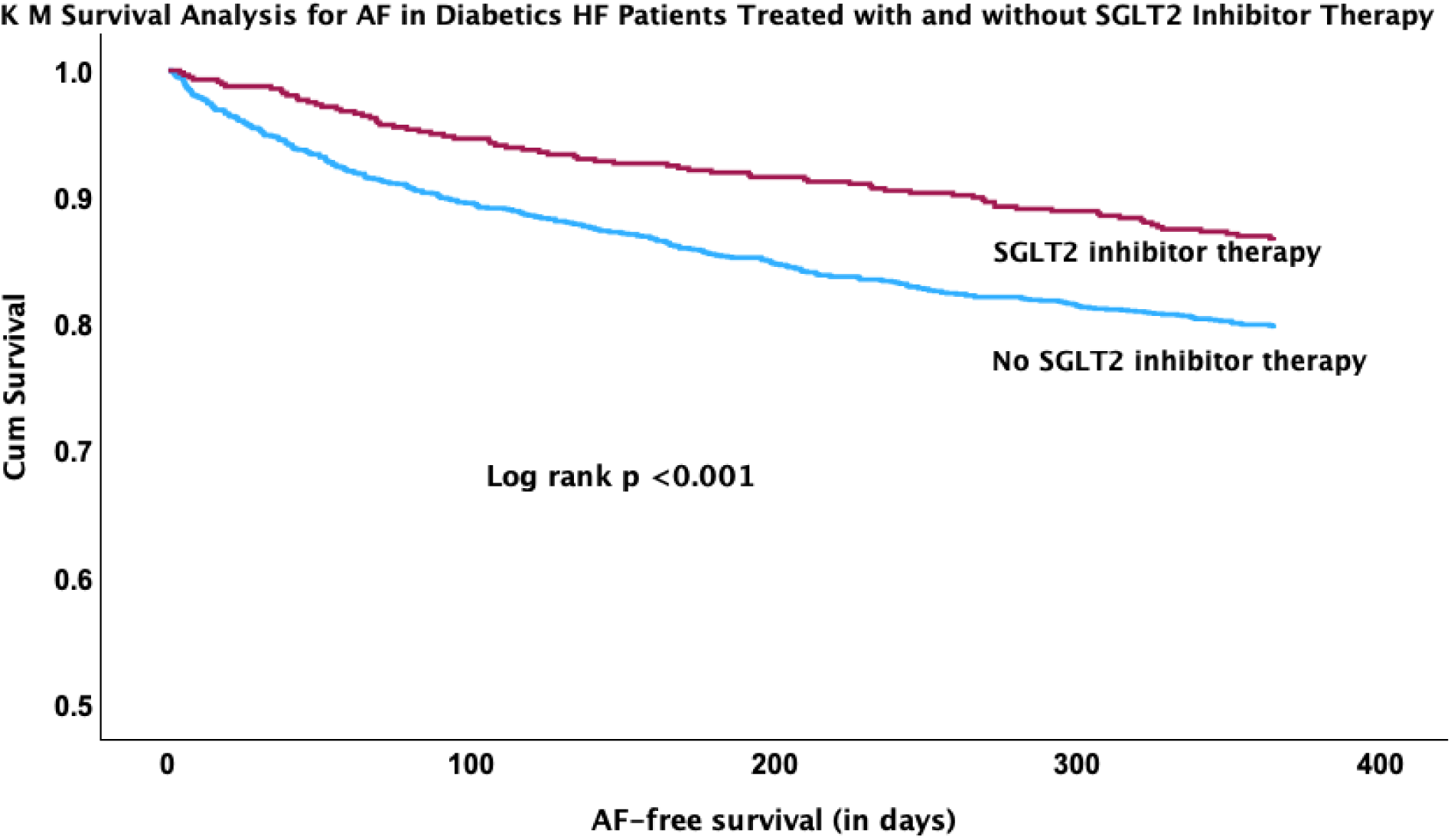
Kaplan-Meier Analysis of the Freedom from AF Following HF Hospitalization for in diabetic patients with and without SGLT2is. Kaplan-Meier analysis of the freedom from AF recurrence after the hospitalization for HFrEF based on SGLT2i therapy. AF = atrial fibrillation; SGLT2i = sodium-glucose cotransporter-2 inhibitor.

**Figure 5.**
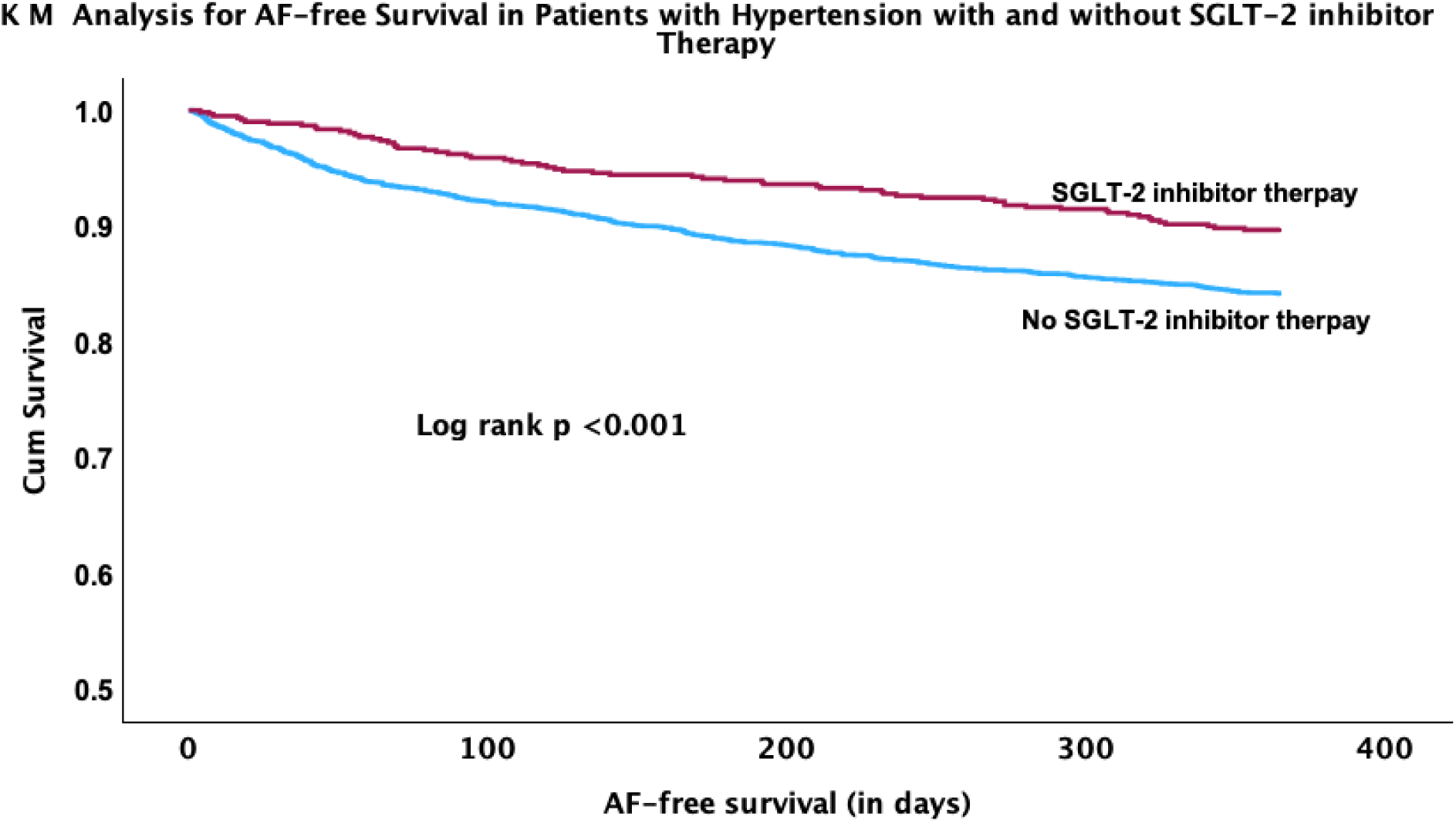
Kaplan-Meier Analysis of HF patients with and without hypertension. Kaplan-Meier analysis of the freedom from AF recurrence after the HF hospitalization based on history of chronic kidney disease. SGLT2i therapy. AF = atrial fibrillation; SGLT2i = sodium-glucose cotransporter-2 inhibitor

**Figure 6.**
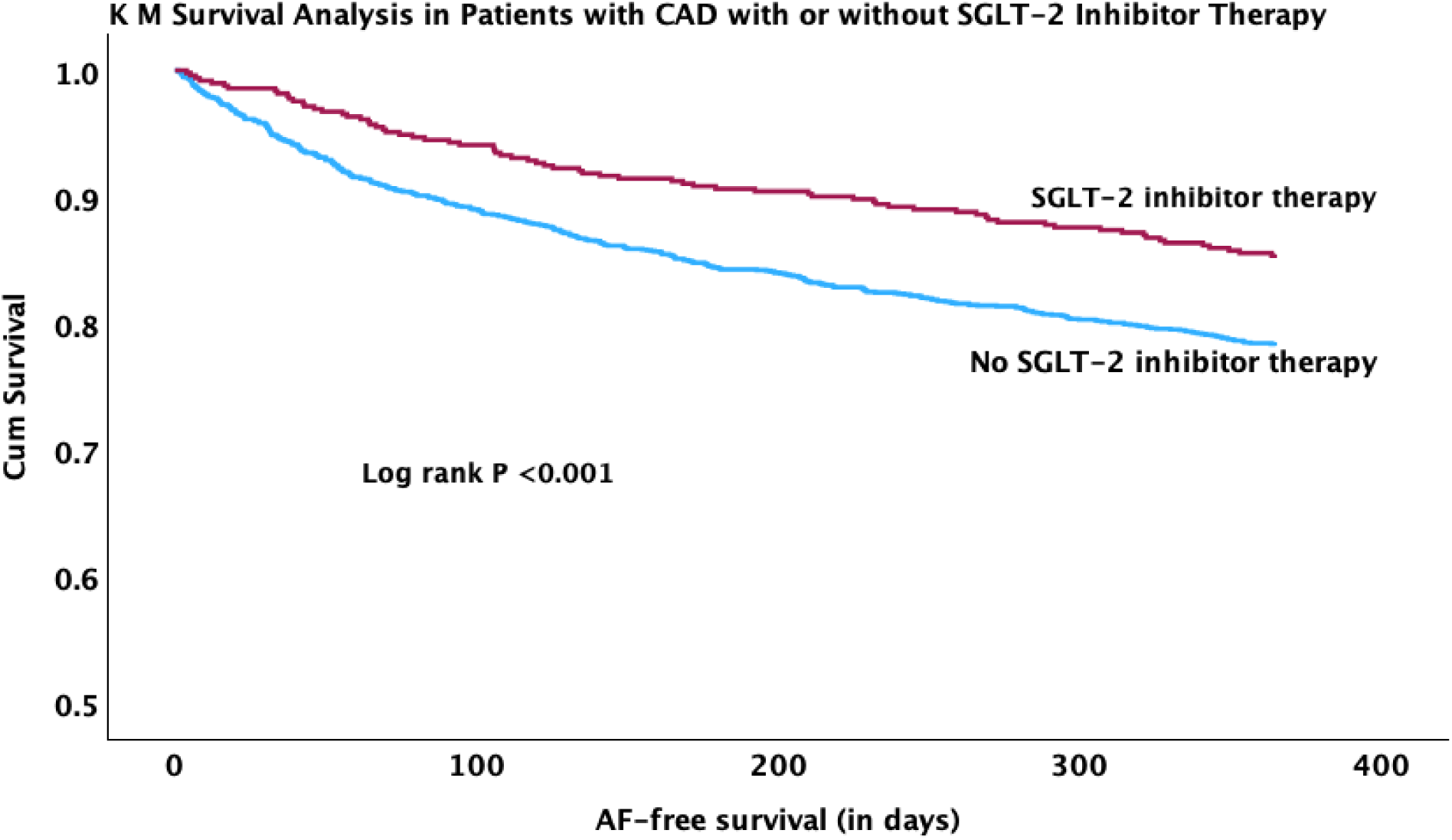
Kaplan-Meier Analysis of the Freedom from AF Following HF Hospitalization for in patients with CAD and, with and without SGLT2is. Kaplan-Meier analysis of the freedom from AF recurrence after the hospitalization for HFrEF based on SGLT2i therapy. AF = atrial fibrillation; SGLT2i = sodium-glucose cotransporter-2 inhibitor.

**Figure 7.**
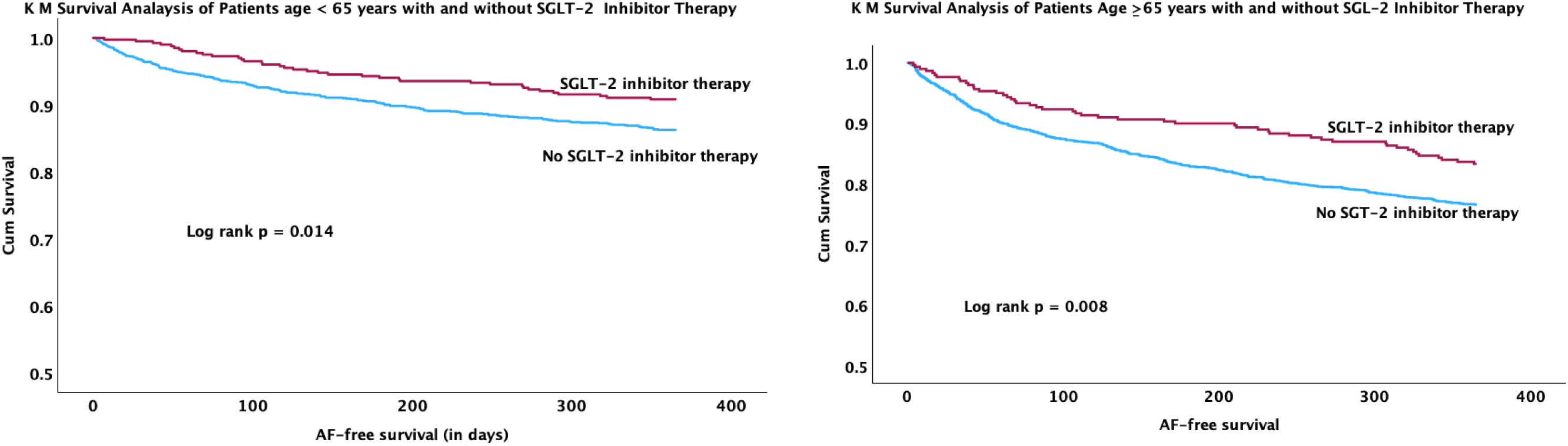
Kaplan-Meier Analysis of the Freedom from AF Following HF Hospitalization for in patients with age <65 years and ≥65 years and, with and without SGLT2is. Kaplan-Meier analysis of the freedom from AF recurrence after the hospitalization for HFrEF based on SGLT2i therapy. AF = atrial fibrillation; SGLT2i = sodium-glucose cotransporter-2 inhibitor.

**Figure 8.**
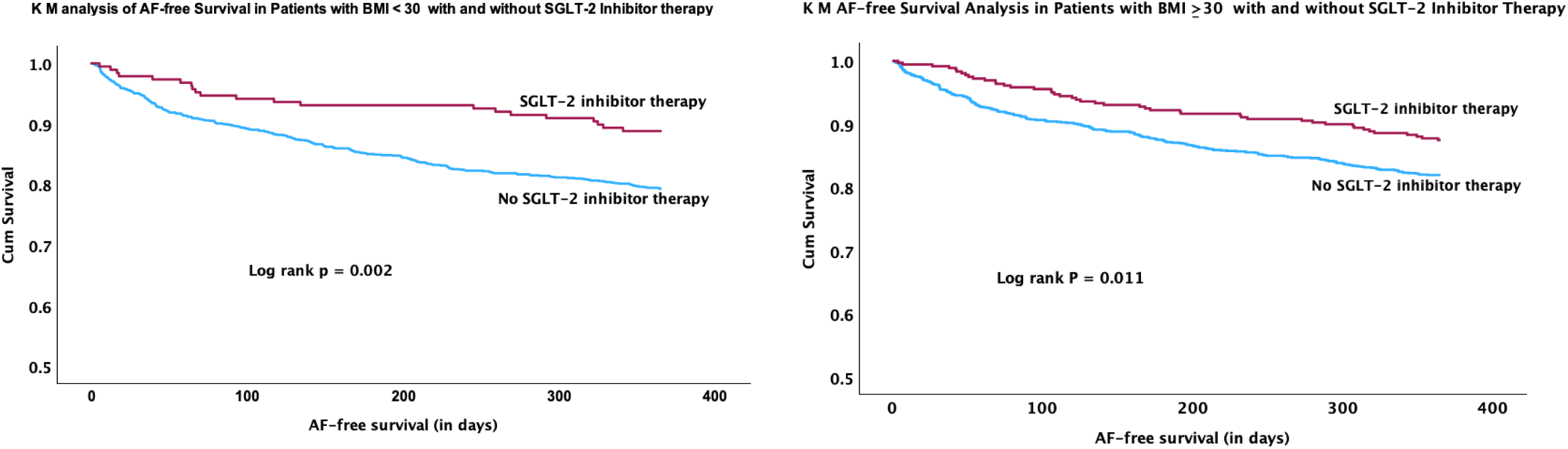
Kaplan-Meier Analysis of the Freedom from AF Following HF Hospitalization for in patients with BMI ≤30 and >30 and, with and without SGLT2is. Kaplan-Meier analysis of the freedom from AF recurrence after the hospitalization for HFrEF based on SGLT2i therapy. AF = atrial fibrillation; BMI= body mass index in Kg/m^2^, SGLT2i = sodium-glucose cotransporter-2 inhibitor.

**Figure 9.**
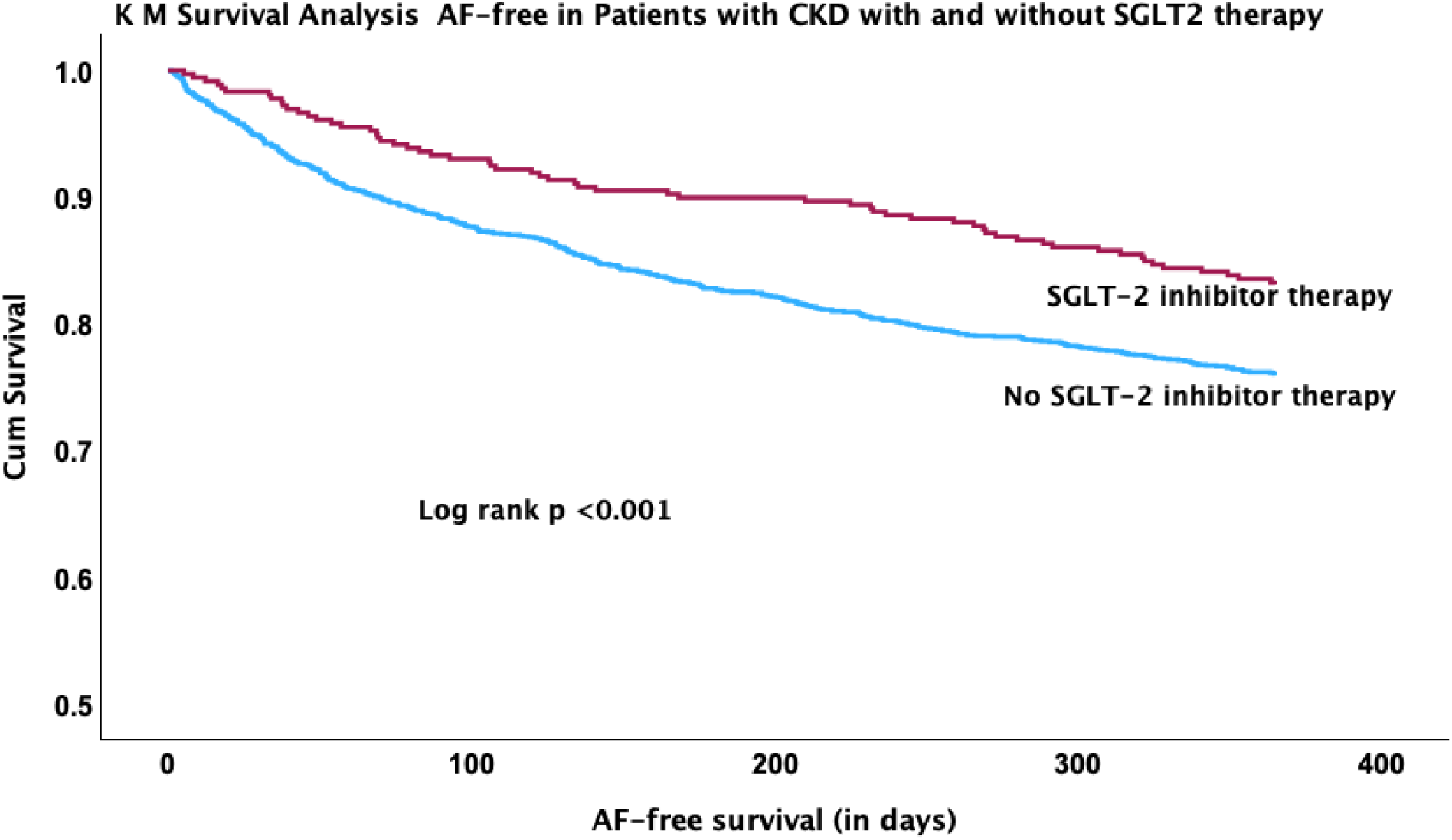
Kaplan-Meier Analysis of HF patients with and without SGLT2i therapy in chronic kidney disease. Kaplan-Meier analysis of the freedom from AF recurrence after the HF hospitalization based on history of chronic kidney disease. SGLT2i therapy. AF = atrial fibrillation; CKD = chronic kidney disease, SGLT2i = sodium-glucose cotransporter-2 inhibitor.

## Discussion

This retrospective study included 3,953 patients hospitalized for HF without a prior diagnosis of AF, of which 18.2% developed AF within 1-year of discharge. Our study demonstrates that: 1) the use of a SGLT2i is associated with a 7.4% absolute and 42% relative risk reduction (12.1% with SGLT2i use vs .19.5% without SGLT2i use) of AF in a 1-year follow up (*HR 0.69, 95% CI: 0.55-0.87; p=0.002*); 2) among those who developed AF during follow-up, SGLT2i therapy was associated with a significant delay in the onset of AF as compared to patients who did not receive SGLT2i therapy, however it may not be clinically that significant 3) SGLT2i use significantly reduced rate of new-onset AF in patients with risk factors such as diabetes and CAD; and 4) SGLT2i therapy was effective regardless of whether patients were ≥65 years old or <65 years old, and irrespective of whether their BMI was ≥30 or <30 kg/m². These findings were consistent across HF subtypes (HFrEF, HFmrEF and HFpEF). Additionally, patients with a history of chronic kidney disease, COPD, and hyperlipidemia showed a higher incidence of AF during follow-up. To our knowledge, this is the first study to evaluate the efficacy of SGLT2is in preventing new-onset AF following HF hospitalization.

Hospitalization for heart failure (HF) significantly increases mortality compared to those never admitted for HF.^16^ Likewise, the development of AF in patients with HF is common and associated with increased morbidity and mortality. Maintaining sinus rhythm improves functional status in patients with HF^17^ and is associated with decreased cardiovascular events. While some therapies, such as catheter ablation,^18^ have demonstrated mortality and functional benefit for treating AF in patients with HFrEF, it is invasive and with limited efficacy, highlighting the need for novel preventive therapies. Therefore, identifying the role of SGLT2i in HF is significant since reducing the occurrence of new onset AF may have a positive impact in survival. Despite the fact that SGLT2is are recommended by the AHA for the treatment of both reduced and preserved ejection fraction HF, only one in five HF patients are prescribed this therapy.^19^ This underuse may be due to the high cost of SGLT2i. This study, despite its retrospective nature and limitations, suggests that SGLT2is may offer anti-arrhythmic benefits in this vulnerable population.

There is growing clinical evidence for the anti-arrhythmic benefit of SGLT2is.^15^ Two recent metanalyses using prospective randomized clinical trials of SGLT2is demonstrated a 19% relative reduction in AF burden^20^ and a 25% relative reduction in adverse events from AF in diabetics or HF.^10^ These findings suggest SGLT2is may reduce AF incidence or recurrence, with consistent effects across various EFs, diabetic status, and chronic kidney disease. A prospective trial found that SGLT2is, compared to dipeptidyl peptidase-4 inhibitors, significantly reduced AF recurrence one year after catheter ablation in diabetic patients. Other retrospective studies are consistent with these results. Abu-Qaoud et al^21^ examined the impact of SGLT2is on AF recurrence following catheter ablation in patients with DM. They demonstrated a significantly lower risk of cardioversion, new anti-arrhythmic medications, and redo AF ablation in patients taking SGLT2is at 12 months following the procedure. Two retrospective studies examined SGLT2i therapy in patients with cancer-therapy-associated cardiac dysfunction also suggest arrhythmia benefit. Gongora et al^22^ demonstrated a reduction in their primary outcome, a composite of HF incidence and hospitalization, and clinically significant arrhythmias, while Avula et al^23^ presented a significant decrease in incidence of AF as a secondary outcome. Though these studies are retrospective and share similar limitations as this study, the results presented are consistent with previous results and add to the evidence for anti-arrhythmic benefit in other clinical settings.

The exact mechanism by which SGLT2is confer anti-arrhythmic benefit is not fully understood and is likely multifactorial. SGLT2i therapy is associated with reduced oxidative stress and chronic inflammation, which may offer a protective effect on AF. ^24^ Studies suggest that, beyond glucose control, SGLT2i have additional metabolic effect on circulating metabolites with a reduced risk of AF. The total concentration of lipoprotein particles and particularly the concentration of HDL particles might mediate this association given the anti-inflammatory and antioxidant properties of HDL.^25^ SGLT2i results in alteration in lipid metabolism and HDL may have a protective role on AF occurrence. ^26^ At cellular and molecular level, SGLT2is have shown to downregulate of CaMK II activity, inhibition of NHE-1, repair of Ca^2+^ handling, stabilization of Na^+^ imbalances, by reducing cardiac load, improving myocardial energy metabolism, inhibiting inflammation, improving myocardial remodeling, reducing sympathetic nerve activity, weight loss and renal function modifications to exert antiarrhythmic effects.^27^ Furthermore, in-vitro studies include improved hemodynamics effects of SGLT2i through natriuresis and glucosuria, improved cardiac metabolism, and reduction of inflammation leading to decreased cardiac fibrosis.^28,29^ Both direct effects of SGLT2is on the myocardium and systemic benefits contribute to the cardioprotective effects demonstrated by this class of medications.

SGLT2is are generally well-tolerated and have a favorable safety profile, though they are associated with an increased risk of urinary tract infections.^30^ Canagliflozin may be associated with increased risk of lower extremity amputations, but this has not been seen in real world data of SGLT2i use and the FDA recently removed this black-box warning.^31^ It is generally accepted that the cardiac and renal benefits largely outweigh risks associated with SGLT2is.^32^ SGLT2is have demonstrated benefit for patients with HF, CKD, and diabetes in prospective trials. These therapies, along with GLP-1 agonists^33,34^ have brought and increased focus to cardiometabolic disease and is a promising area of further research.

### Study limitations

As a retrospective analysis, this study has several limitations. First, the data used in this study was extracted from an electronic medical record within a single healthcare, relying on the database’s accuracy. Due to the nature of the record system, events captured outside of the health system would not be reflected in this study unless accurately recorded in subsequent encounters. To mitigate this bias, we ensured each patients had at least two encounters with this health care system. Though, the use outpatient encounter within the health system to define an endpoint for our control group introduces significant bias. This increases the chances of capturing AF diagnoses in this group reducing the difference in incidence seen between groups, but acts in the opposite direction as well, increasing the accuracy and incidence of SGLT2i prescription. This study was also underpowered to stratify patients by specific agents or dose of SGLT2i. Like all retrospective studies, ours also has a potential selection bias attributable to unmeasured factors, despite controlling for these variables prior to analysis.

## Conclusions

Our analysis suggests that, among patients hospitalized with a primary diagnosis of HF with no prior diagnosis of AF, treatment with SGLT2i is associated with a significantly lower incidence of AF during a one-year follow-up. This effect is consistent across HF subgroups (HFpEF, HFmEF and HFrEF) and in the presence of hypertension, diabetes and CKD. SGLT2i use in HF also delay the development of AF. Larger, prospective randomized controlled studies are necessary to confirm these findings.

## Data Availability

All data was collected by authors and available

## Funding Support and Author Disclosures

We recognize and appreciate IU Health for their assistance and support in this study. Dr. Mithilesh Das is supported by [NIH-R01 HL158952-01A1) grant. All authors have reported that they have no relationships relevant to the contents of this paper to disclose.

## Perspectives

In patients with hospitalization for HF with no history of AF, treatment with SGLT2i therapy reduces the risk development of AF. SGLT2i therapy should be considered in the management of patients with AF in addition to traditional AF treatments.

## Translational outlook

Transitional studies are needed to further evaluate the mechanisms of SGLT2i in cardiac remodeling and preventing AF.

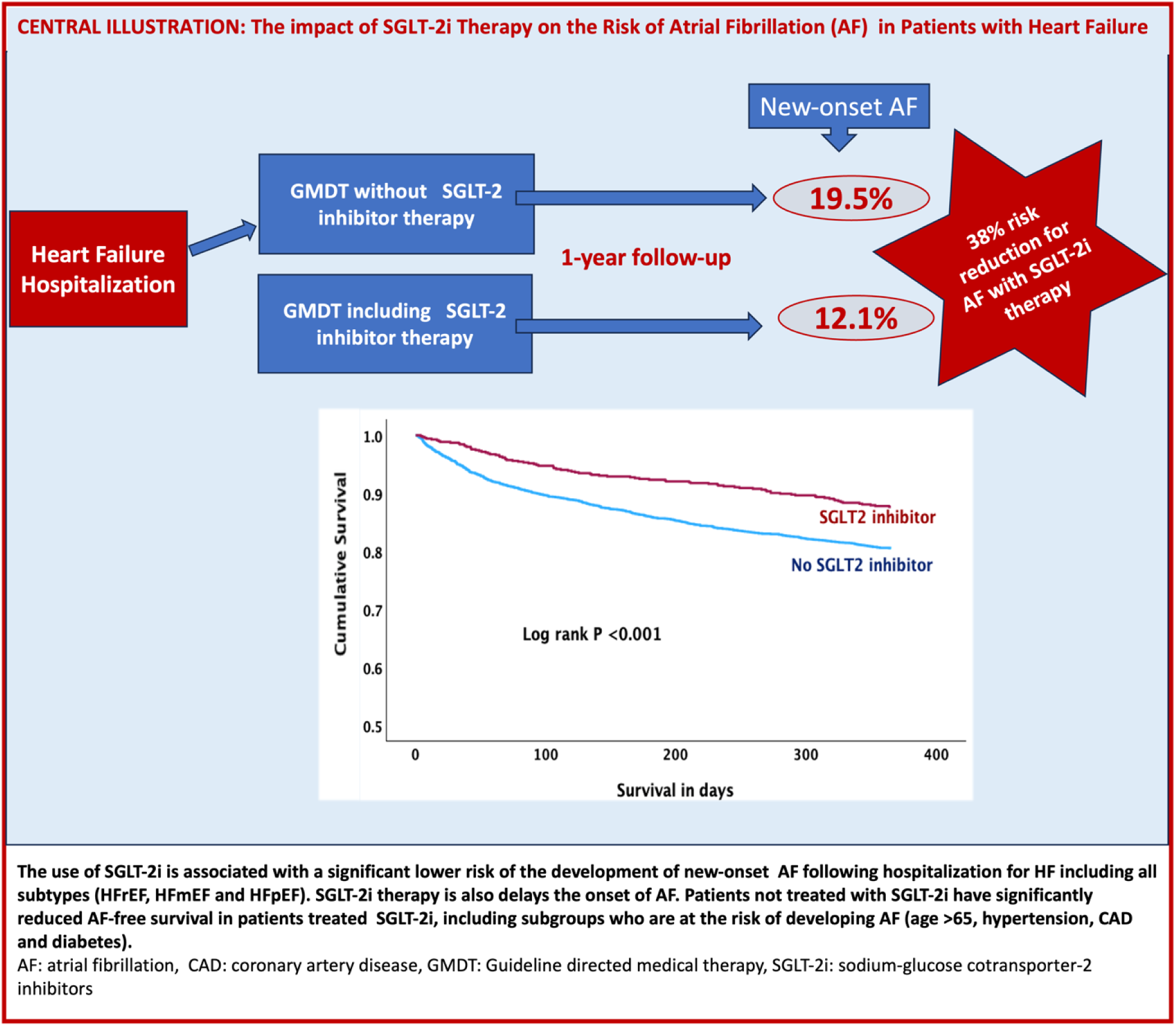

## Notes

### Competing Interest Statement

The authors have declared no competing interest.

### Funding Statement

NIH grant: NIH-R01 HL158952-01A1

### Author Declarations

Exemption from Institutional Review Board (IRB) approval was obtained from the Indiana University given the deidentified nature of the data

